# Impact of WHO AWaRe Antibiotic Handbook training on antibiotics prescribing knowledge among primary care providers: A vignette-based, pre-post pilot study in Patna, India

**DOI:** 10.1101/2025.09.06.25335195

**Authors:** Poshan Thapa, Prachi Shukla, Chandrashekhar Joshi, Sena Sayood, Pradeep Kumar Sinha, Diwash Timilsina, Mili Dutta, Madhukar Pai, Samira Abbasgholizadeh Rahimi, Sumanth Gandra

## Abstract

**Introduction:** Inappropriate antibiotic prescribing is a major concern in low– and middle-income countries (LMICs), particularly at the primary care level. The WHO AWaRe Antibiotic Handbook was introduced to promote rational antibiotic use, yet its real-world feasibility and potential impact remain underexplored. Our study evaluated the effectiveness and usefulness of the WHO AWaRe Handbook training among primary care providers (PCPs) in Patna, India.

**Methods:** We conducted a pre-post interventional study among 145 PCPs (40 formal providers (FPs) and 105 informal providers (IPs), 98% male) in Patna, India. Participants received training from an infectious disease physician on the WHO AWaRe Antibiotic Handbook. Antibiotic prescribing knowledge was assessed before and after the intervention using clinical vignettes for four conditions: acute diarrhea, urinary tract infection (UTI), cellulitis, and community-acquired pneumonia (CAP). An endline survey evaluated the perceived usefulness of the training. Changes in prescribing were analyzed using McNemar’s test for paired data.

**Results:** The intervention significantly reduced overall antibiotic prescribing for acute diarrhea (p=0.0003) and UTI (p=0.0113), with greater reductions among IPs. No significant changes were observed for cellulitis (p=0.3692) or CAP (p=0.7150). Watch-category antibiotic prescribing significantly decreased for acute diarrhea (p<0.0001), with no significant changes for other conditions. IPs showed greater improvements overall compared to FPs. The majority of providers (75%; n=107) rated the training as moderately or very useful.

**Conclusion:** Training PCPs using the WHO AWaRe Handbook improved antibiotic prescribing knowledge for some common conditions, particularly among IPs. Future research should focus on the impact of ongoing training, tailored interventions, and long-term follow-up.

**Strengths and limitations of this study:**

– This is the first study to evaluate the effectiveness of WHO AWaRe Handbook training on improving antibiotic prescribing knowledge among primary care providers in India, focusing on both formal and informal healthcare providers.
– Using a vignette-based, pre-post study design allowed for standardized assessment of prescribing knowledge across four common clinical conditions: acute diarrhea, cellulitis, pneumonia, and urinary tract infection.
– Stratified analysis by provider type offered important insights into the intervention’s differential effects, particularly highlighting knowledge improvements among informal providers.
– While the study captures shifts in prescribing knowledge, it does not assess actual prescribing behavior in clinical practice, which may limit the generalizability of the findings.
– The study evaluated outcomes over a short follow-up period, which restricts understanding of the sustainability of training effects over time.

## Introduction

Global antibiotic use increased 16.3% (2016–2023), mainly in low– and middle-income Countries (LMICs) (+18.6%), while high-income countries saw a 4.9% decline. ^1^ Overuse and misuse of antibiotics remain a significant challenge, especially at the primary care level, where over 50% of outpatient visits in LMICs result in an antibiotic prescription, often without clinical justification. ^2^ Standardized patient studies have shown high rates of inappropriate prescribing, with broad-spectrum antibiotics excessively used in China and India. ^3^ The COVID-19 pandemic further exacerbated antibiotic overuse, with India’s antibiotic consumption increasing by 103% from 2000 to 2015, making it the largest consumer of antibiotics globally.^4^ The challenge of antibiotic over-consumption is particularly complex in India due to its fragmented healthcare system, which includes a range of formal and informal providers, many of whom prescribe antibiotics empirically, often without regulatory oversight. ^3^ ^5^

In India, primary health care within the government’s formal system is predominantly provided through Primary Health Centres (PHCs) and their subsidiary Sub-Centres (SCs). ^6^ However, India’s health system also includes a large private sector, which provides approximately 87% of initial primary care, particularly in underserved regions. The private health sector comprises a spectrum of providers, from small clinics or practitioner-run setups to multispecialty hospitals. At the primary care level, providers vary significantly in qualifications, ranging from highly qualified formal providers and specialists to informal healthcare providers (IPs). ^7^ ^8^ Formal private providers typically hold Bachelor of Medicine, Bachelor of Surgery (MBBS) degrees, whereas IPs, commonly called rural medical practitioners or village doctors, operate outside the formal health system and lack accredited medical qualifications. Despite this, they frequently dispense allopathic treatments, including antibiotics and injections, without formal training. ^8–10^ Additionally, practitioners of Ayurveda, Yoga & Naturopathy, Unani, Siddha, and Homeopathy (AYUSH) are known to prescribe and dispense antibiotics in India ^11^, despite not being legally authorized to prescribe modern medicine, including antibiotics.

Considering the need for global antibiotic stewardship, WHO introduced the AWaRe (Access, Watch, Reserve) classification system in 2017. This framework emphasizes the preferential use of narrow-spectrum “Access” antibiotics over broad-spectrum “Watch” and last-resort “Reserve” antibiotics, with WHO recommending that at least 60% of all antibiotic use should consist of Access-group antibiotics. ^12^ In November 2022, WHO expanded this initiative by publishing the AWaRe Antibiotic Handbook, a clinical decision-support tool providing evidence-based guidance on antibiotic necessity, choice, dosage, route, and duration. ^13^

Although the AWaRe Handbook is a promising tool for antimicrobial stewardship (AMS), its adoption in LMICs faces challenges. Studies indicate that clinical decision-support tools are often underutilized unless adapted to local contexts and effectively delivered to primary care providers (PCPs). ^14^ ^15^ To address this, WHO published the AWaRe Handbook under a Creative Commons (CC) license, enabling local adaptations, language translations, and simplified decision aids. Early evidence from China suggests a traffic-light clinical guideline approach reduced antibiotic prescribing from 82% to 40% in the intervention group, compared to a 5-percentage-point reduction in the control group. ^16^ However, similar research is lacking in India, where over-prescription of broad-spectrum antibiotics remains prevalent. Furthermore, translation into regional languages and integration into existing clinical workflows may be necessary to enhance uptake among PCPs, especially IPs, who may have limited access to standardized treatment guidelines.

Therefore, we conducted a pilot study using the WHO AWaRe Handbook to evaluate the impact of training PCPs in Patna, India. Our findings contribute to the growing evidence on AMS interventions in LMICs and inform efforts to scale up AWaRe Handbook implementation in primary care settings, particularly within the informal sector, where evidence remains limited.

## Methods

### Design

We conducted a pre-post interventional study in Patna, India, to assess changes in antibiotic prescribing knowledge before and after a training intervention. Similar approaches have been used in studies with comparable objectives. ^17–19^ The study was conducted in collaboration with World Health Partners (WHP), a local non-governmental organization (NGO) with over a decade of experience working with the private healthcare sector across multiple districts in various states and districts in India, including Patna.

### Study site

Our study was conducted in Patna, Bihar’s capital and largest city, which serves as the district’s administrative headquarters. As per the 2011 census, the urban agglomeration had a population of 2.05 million. ^20^ Patna was selected due to WHP’s ongoing work and collaborations, which facilitated field implementation and participant enrollment, making it an operationally feasible location.

### Study population and sampling

The study included private primary care providers (PCPs) offering consultation services in independent clinics across Patna. Both formal providers (FPs) and IPs were enrolled. AYUSH providers were pragmatically grouped with IPs to align with prescribing regulations in India, as neither group is authorized to prescribe modern medicine, including antibiotics. Convenience sampling was used to recruit providers, leveraging WHP’s existing network across 75 wards of Patna city. WHP field officers, who routinely engage with private providers for various project activities, compiled a list of eligible participants. For IPs, snowball sampling was also employed, with enrolled providers referring eligible peers. Providers were invited through in-person visits or phone calls, and those providing written informed consent were enrolled. Of the 325 providers approached, 146 (45%) consented to participate, including 40 FPs and 106 IPs (8 AYUSH and 98 IPs).

### Sample Size

As this was a pilot study, the sample size was based on feasibility and available resources. The final sample of 145 providers aligns with recommendations suggesting 100–120 participants for pilot studies to estimate key parameters. ^21^ The use of a pre-post paired design further reduced the variability. While sufficient for exploratory analysis, the findings should be interpreted as preliminary to guide future larger-scale studies.

### Description of intervention

All participating providers received training on the WHO AWaRe Antibiotic Handbook, delivered by a US board-certified infectious disease physician (SG) who was also trained in India and contributed to the development of the Handbook. Training sessions were conducted separately for FPs and IPs, typically lasting 3 to 4 hours each. The training covered an overview of antibiotics, antimicrobial resistance, rational prescribing, the AWaRe classification, and its application. Case-based discussions were conducted for four common conditions associated with widespread antibiotic use: acute diarrhea, acute pharyngitis, community-acquired pneumonia (CAP), and lower urinary tract infection (UTI).

Participants were encouraged to ask questions, and discussions addressed field-level prescribing challenges and AWaRe guidelines. Providers were also introduced to the AWaRe Handbook’s Mobile and Web Application, assisted in downloading it, and provided a tutorial on its use. At the end of the training, FPs received a hard copy of the AWaRe Handbook (English), and IPs received Hindi-translated infographics covering eight conditions: acute bronchitis, acute diarrhea, acute otitis media, acute pharyngitis, acute sinusitis, cellulitis, CAP and UTI. The training was conducted in two phases: 53 attendees in December 2023 and 93 attendees in March 2024 at locations selected for accessibility and participation. To ensure consistency in this study’s clinical vignettes, the term “prescribing” was used for both FPs and IPs, though IPs typically “dispense” rather than “prescribe” antibiotics.

### Outcome and measurement

Provider knowledge was assessed using clinical vignettes, a well-established method for evaluating changes in provider knowledge and clinical decision-making in antibiotics research. ^22–24^ The vignettes were developed through discussions among co-authors, led by infectious disease physicians (SG and SS), and adapted from validated vignettes used in previous studies. ^25^ ^26^ The final set of vignettes (Table 1) was pilot tested with ten providers, and feedback indicated that providers understood the case scenarios well. Initially, eight vignettes were planned, but due to provider concerns over completion time, the number was reduced to four: acute diarrhea, cellulitis, CAP (mild severity), and UTI. A small proportion (∼5%) of providers completed only two or three vignettes instead of all four.

**Table 1:**
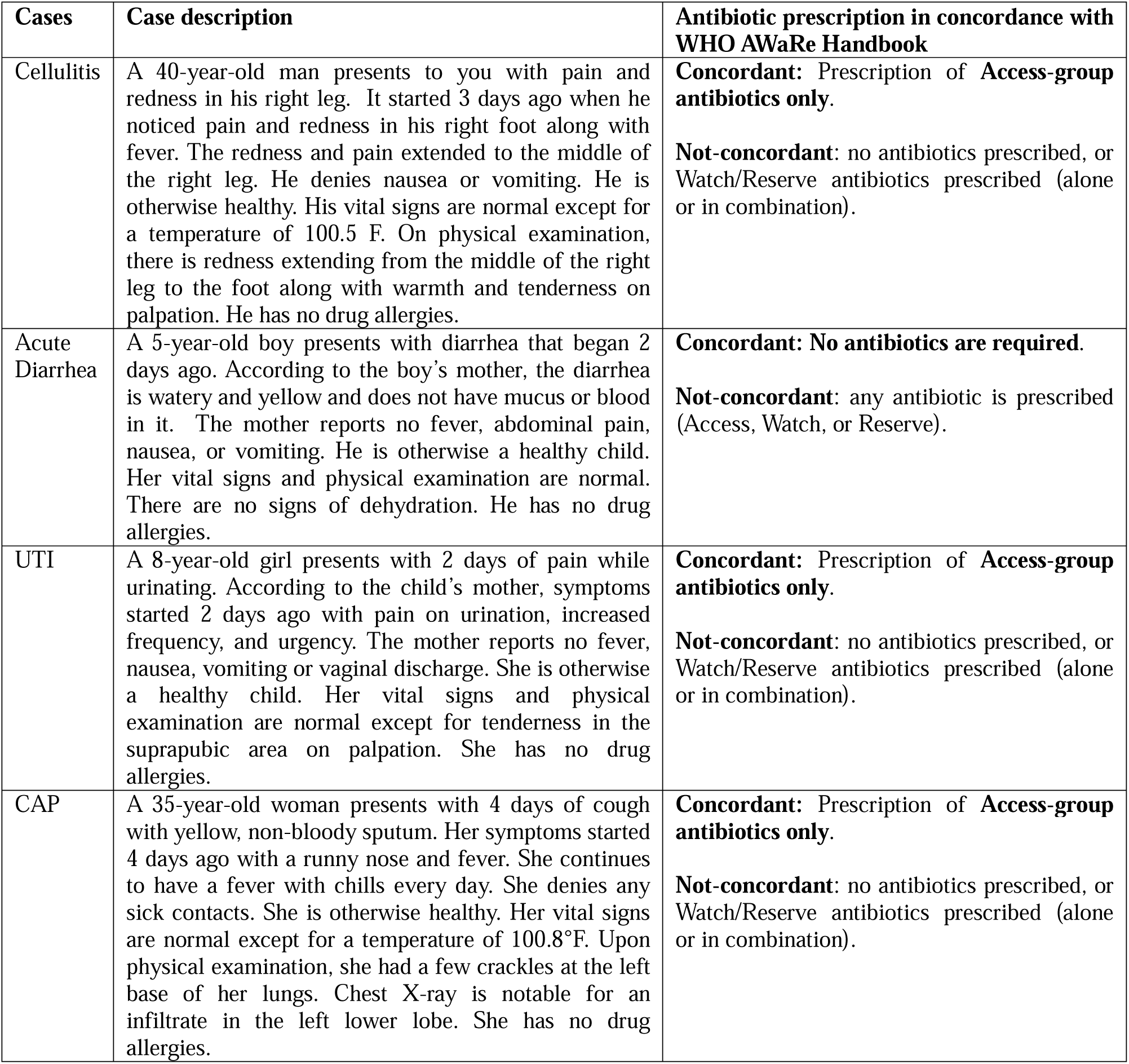
Vignette cases and antibiotic prescription in concordance with the WHO AWaRe Antibiotic Handbook.

Since the primary aim of this study was not to assess case management accuracy (such as history taking, laboratory workup and final diagnosis), results are presented based on changes and shifts in antibiotic prescribing patterns for the four vignette conditions before and after the intervention. The primary outcome was the change in overall antibiotic prescriptions and Watch-category antibiotic use for each vignette condition. The secondary outcome measures were changes in the proportion of antibiotic prescriptions and the concordance of antibiotic prescriptions per the AWaRe Handbook for each clinical condition, as described in Table 1.

During data collection, interviewers first recorded the participants’ socio-demographic information. Providers then completed the vignettes in printed paper format, which were self-administered unless assistance was requested from field officers, which occurred in a few cases. The vignettes were primarily administered at provider clinics by WHP field officers, who received training beforehand. A subset of providers preferred to complete the vignettes before the training sessions at the training site; in such cases, providers were seated separately to minimize response contamination.

Following the intervention, WHP field officers conducted follow-up vignettes at provider clinics after 2–3 months to allow providers to familiarize themselves with the AWaRe Handbook and mobile application and integrate the tool into their daily practice. In addition to the vignettes, providers were interviewed using a semi-structured questionnaire to gather feedback on the intervention. The baseline and follow-up data collection took an average of 30 to 40 minutes per provider.

### Data preparation and analysis

Vignette data collected on paper forms were entered into a Microsoft Excel-based system, designed and tested with 5% of the dataset. Data entry was conducted at the clinical condition level rather than the provider level, as most providers completed up to four vignettes. Consequently, analyses were based on case numbers rather than provider numbers. A co-author with a medical background cross-checked and verified antibiotic prescriptions, followed by AWaRe framework classification, with a second verification carried out by another co-author. The dataset was then imported into Stata (version 18, StataCorp, USA) for cleaning and analysis.

Descriptive statistics were used to summarize provider characteristics, with categorical variables reported as frequencies and percentages. Changes in antibiotic prescribing knowledge before and after the intervention were assessed using McNemar’s test for paired binary data. Analyses were stratified by provider type (formal vs. informal providers) and clinical condition (acute diarrhea, cellulitis, CAP, and UTI). The primary analysis assessed changes in overall antibiotic prescription and Watch-category antibiotic use. The secondary analysis examined changes in correct antibiotic prescription according to WHO AWaRe Handbook recommendations. The perceived usefulness of the intervention was evaluated through a semi-structured survey, with responses summarized as frequencies and percentages. Qualitative data were extracted from open-ended questionnaire responses. A descriptive analysis was conducted, and representative quotes were selected to illustrate them.

### Patient and public involvement

Patients and members of the public were not involved in the design, conduct, reporting, or dissemination plans of this research. The study focused on PCPs and assessed changes in antibiotic prescribing knowledge using clinical vignettes. Given the nature of the intervention and the study population, direct patient or public involvement was not deemed feasible or applicable.

## Results

Out of the total 146 consenting providers, 145 (99%) providers attended the training and completed the baseline and endline surveys (pre– and post-vignettes), with 40 FPs and 105 IPs. The endline survey questionnaire, which evaluated the usefulness of the intervention, was completed by 143 providers, as two declined to participate.

### Characteristics of the study population

The median age was 46.5 (24-87) years for FPs and 40 years (18-87) for IPs. Both groups predominantly consisted of male participants, with 98% among FPs and 98% among IPs. The median duration of practice was 15 years for both groups. All FPs were either MBBS (72.5%) or Doctor of Medicine (MD) (27.5%) degree holders, while most IPs (80%) had higher education, and a small proportion (6.5%) held degrees in AYUSH [Table 2].

**Table 2:**
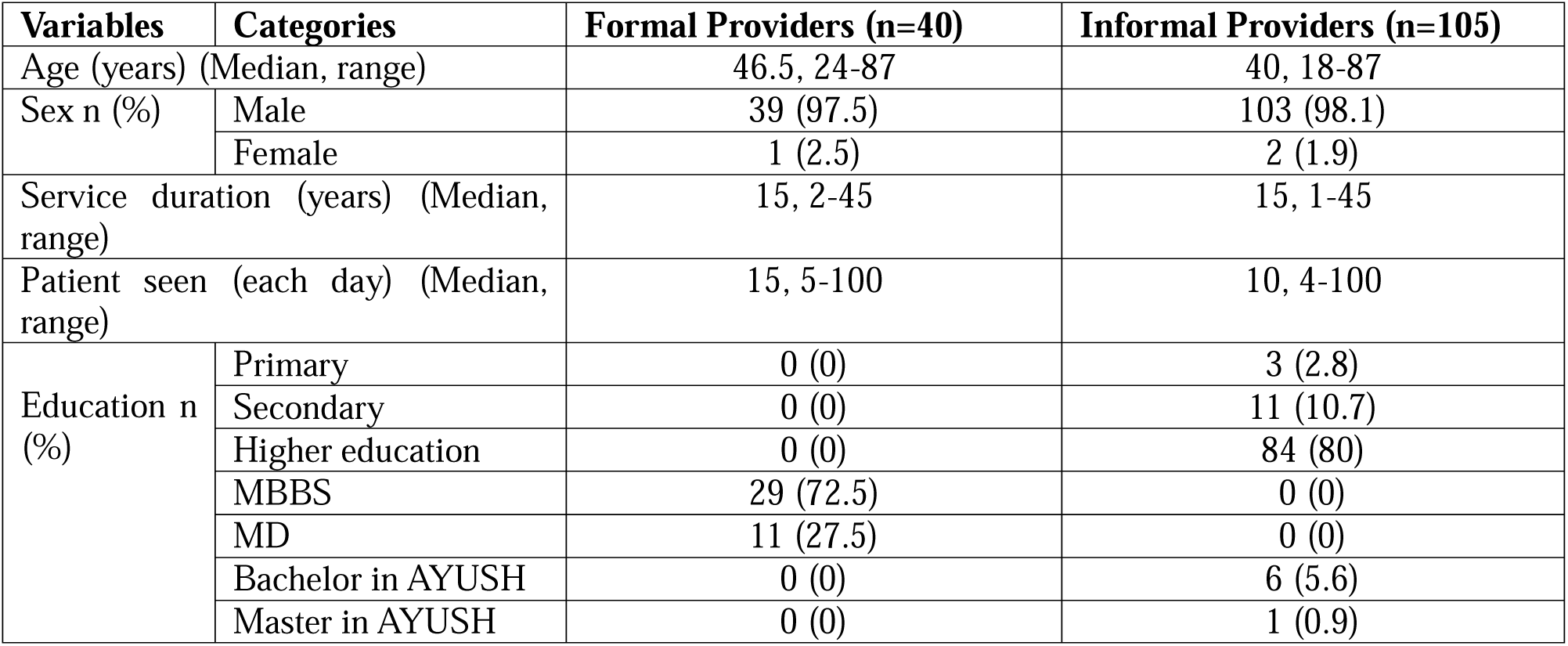
Distribution of the characteristics of the providers (n=145)

### Change in overall antibiotic prescription and Watch-category antibiotic prescriptions for four infectious clinical vignettes before and after the intervention

There were significant changes in antibiotic prescribing following the intervention among acute diarrhea and UTI cases [Table 3]. For acute diarrhea cases (n=145), post-intervention, 23 providers stopped prescribing antibiotics while four started prescribing, resulting in a significant net reduction (p=0.0003). Similarly, for UTI cases (n=144), post-intervention, 31 providers stopped prescribing antibiotics while 14 started prescribing antibiotics (p=0.0113). No significant changes were found for cellulitis (p=0.3692) or CAP (p=0.7150).

**Table 3:** Change in overall antibiotic prescription and Watch-category antibiotic prescriptions for four infectious clinical vignettes before and after the intervention:

| Vignette conditions | Pre-intervention | Post-intervention |  | p-value** |
| --- | --- | --- | --- | --- |
| Overall antibiotic prescription |  |  |  |  |
|  |  | Prescription | No prescription |  |
| Acute Diarrhea (n=145) | Prescription | 79 | 23 | 0.0003* |
|  | No prescription | 4 | 39 |  |
| Cellulitis (n=145) | Prescription | 91 | 18 | 0.3692 |
|  | No prescription | 13 | 23 |  |
| CAP (n=144) | Prescription | 103 | 16 | 0.7150 |
|  | No prescription | 14 | 11 |  |
| UTI (n=144) | Prescription | 59 | 31 | 0.0113* |
|  | No prescription | 14 | 40 |  |
| Overall Watch-category antibiotic prescription<sup>&</sup> |  |  |  |  |
| Acute Diarrhea (n=145) | Prescription | 38 | 37 | 0.0000* |
|  | No prescription | 8 | 62 |  |
| Cellulitis (n=145) | Prescription | 46 | 18 | 0.1404 |
|  | No prescription | 28 | 53 |  |
| CAP (n=144) | Prescription | 33 | 29 | 0.1086 |
|  | No prescription | 18 | 64 |  |
| UTI (n=144) | Prescription | 43 | 24 | 0.1048 |
|  | No prescription | 14 | 63 |  |

\*\*McNemar's test
<sup>&</sup>At least one Watch-category antibiotic (in case of multiple prescriptions)

**Table 4:**
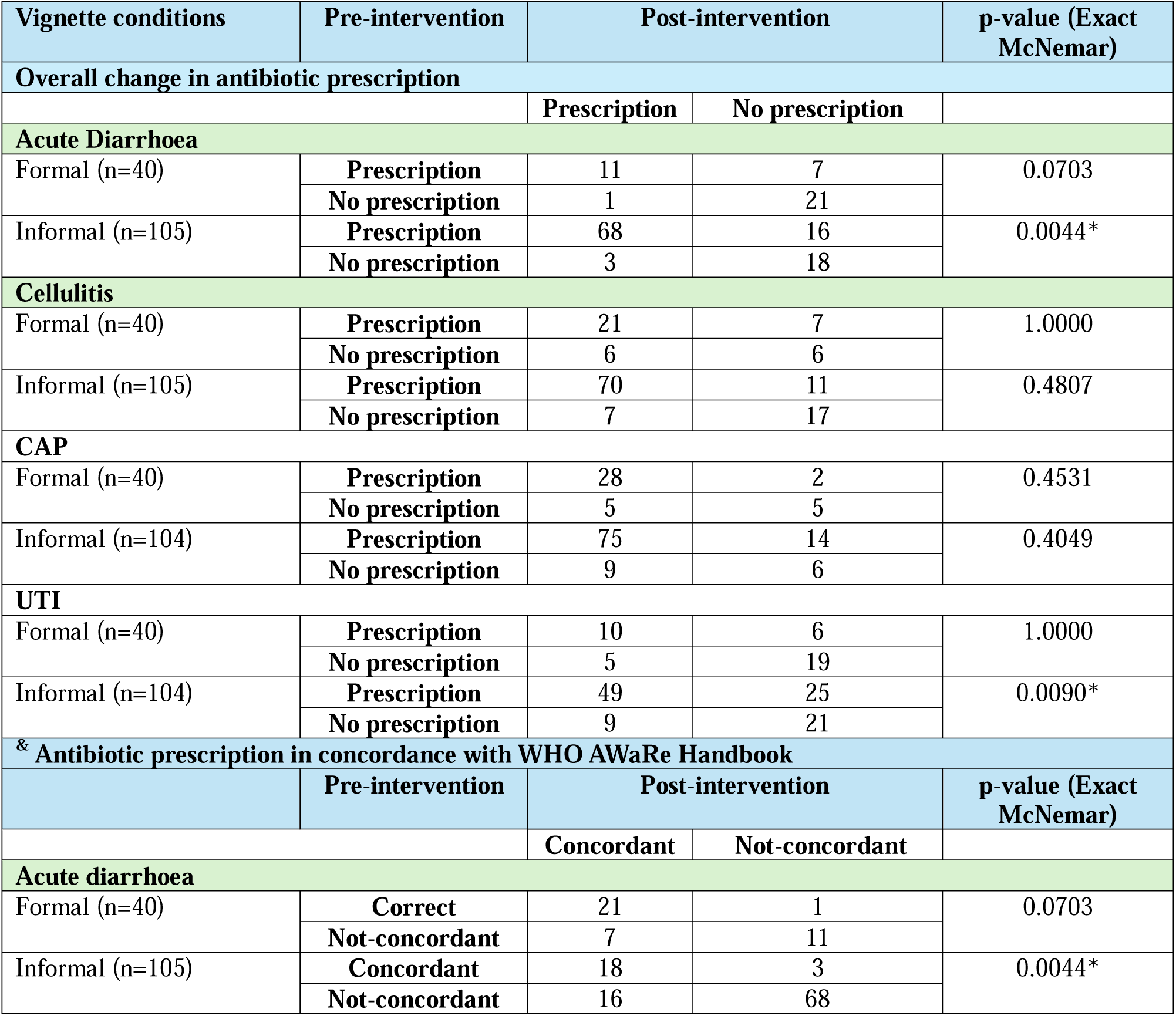
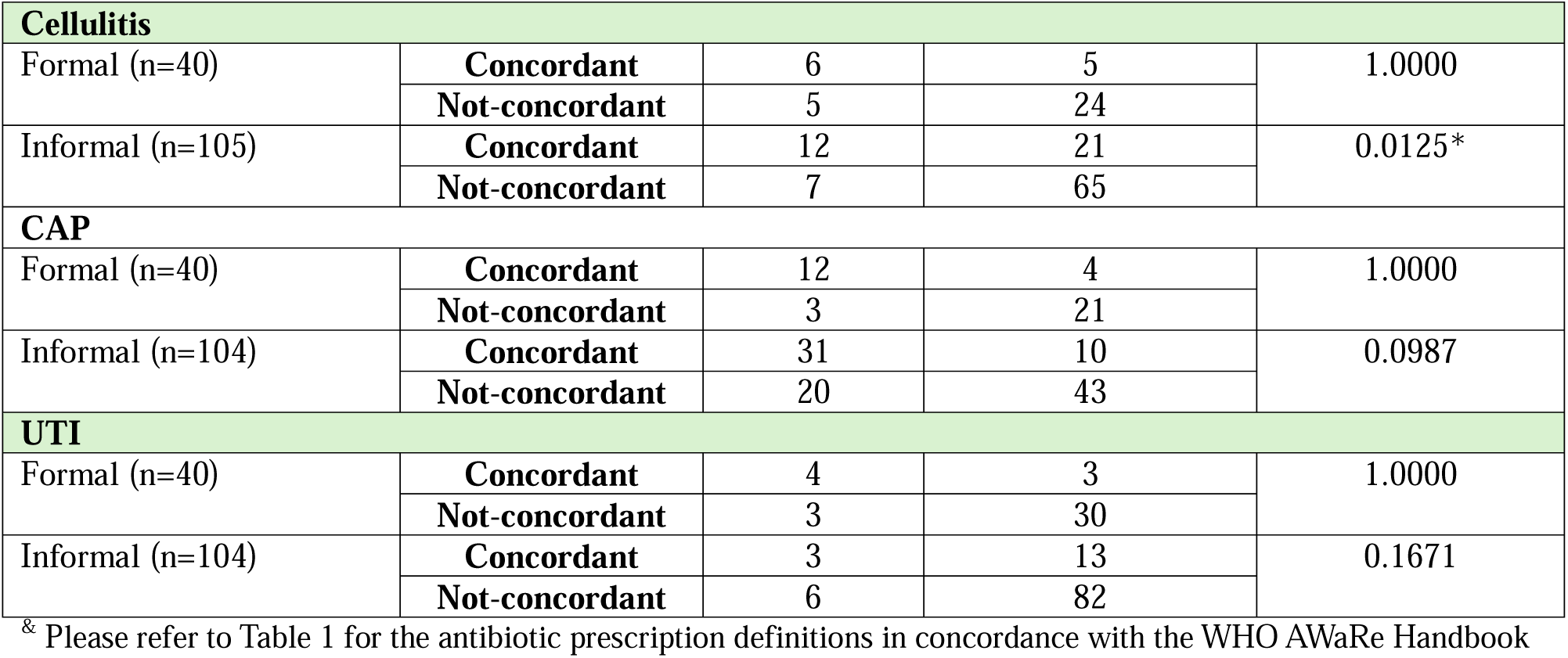
Difference in overall antibiotic prescription and antibiotic prescription in concordance with WHO AWaRe Antibiotic Handbook before and after the intervention among formal and informal providers in Patna, India.

| Vignette conditions | Pre-intervention | Post-intervention |  | p-value (Exact McNemar) |
| --- | --- | --- | --- | --- |
| Overall change in antibiotic prescription |  |  |  |  |
|  |  | Prescription | No prescription |  |
| Acute Diarrhoea |  |  |  |  |
| Formal (n=40) | Prescription | 11 | 7 | 0.0703 |
|  | No prescription | 1 | 21 |  |
| Informal (n=105) | Prescription | 68 | 16 | 0.0044* |
|  | No prescription | 3 | 18 |  |
| Cellulitis |  |  |  |  |
| Formal (n=40) | Prescription | 21 | 7 | 1.0000 |
|  | No prescription | 6 | 6 |  |
| Informal (n=105) | Prescription | 70 | 11 | 0.4807 |
|  | No prescription | 7 | 17 |  |
| CAP |  |  |  |  |
| Formal (n=40) | Prescription | 28 | 2 | 0.4531 |
|  | No prescription | 5 | 5 |  |
| Informal (n=104) | Prescription | 75 | 14 | 0.4049 |
|  | No prescription | 9 | 6 |  |
| UTI |  |  |  |  |
| Formal (n=40) | Prescription | 10 | 6 | 1.0000 |
|  | No prescription | 5 | 19 |  |
| Informal (n=104) | Prescription | 49 | 25 | 0.0090* |
|  | No prescription | 9 | 21 |  |
| & Antibiotic prescription in concordance with WHO AWaRe Handbook |  |  |  |  |
|  | Pre-intervention | Post-intervention |  | p-value (Exact McNemar) |
|  |  | Concordant | Not-concordant |  |
| Acute diarrhoea |  |  |  |  |
| Formal (n=40) | Correct | 21 | 1 | 0.0703 |
|  | Not-concordant | 7 | 11 |  |
| Informal (n=105) | Concordant | 18 | 3 | 0.0044* |
|  | Not-concordant | 16 | 68 |  |

| Cellulitis |  |  |  |  |
| --- | --- | --- | --- | --- |
| Formal (n=40) | Concordant | 6 | 5 | 1.0000 |
|  | Not-concordant | 5 | 24 |  |
| Informal (n=105) | Concordant | 12 | 21 | 0.0125* |
|  | Not-concordant | 7 | 65 |  |
| CAP |  |  |  |  |
| Formal (n=40) | Concordant | 12 | 4 | 1.0000 |
|  | Not-concordant | 3 | 21 |  |
| Informal (n=104) | Concordant | 31 | 10 | 0.0987 |
|  | Not-concordant | 20 | 43 |  |
| UTI |  |  |  |  |
| Formal (n=40) | Concordant | 4 | 3 | 1.0000 |
|  | Not-concordant | 3 | 30 |  |
| Informal (n=104) | Concordant | 3 | 13 | 0.1671 |
|  | Not-concordant | 6 | 82 |  |

\* Please refer to Table 1 for the antibiotic prescription definitions in concordance with the WHO AWaRe Handbook

**Table 4:**
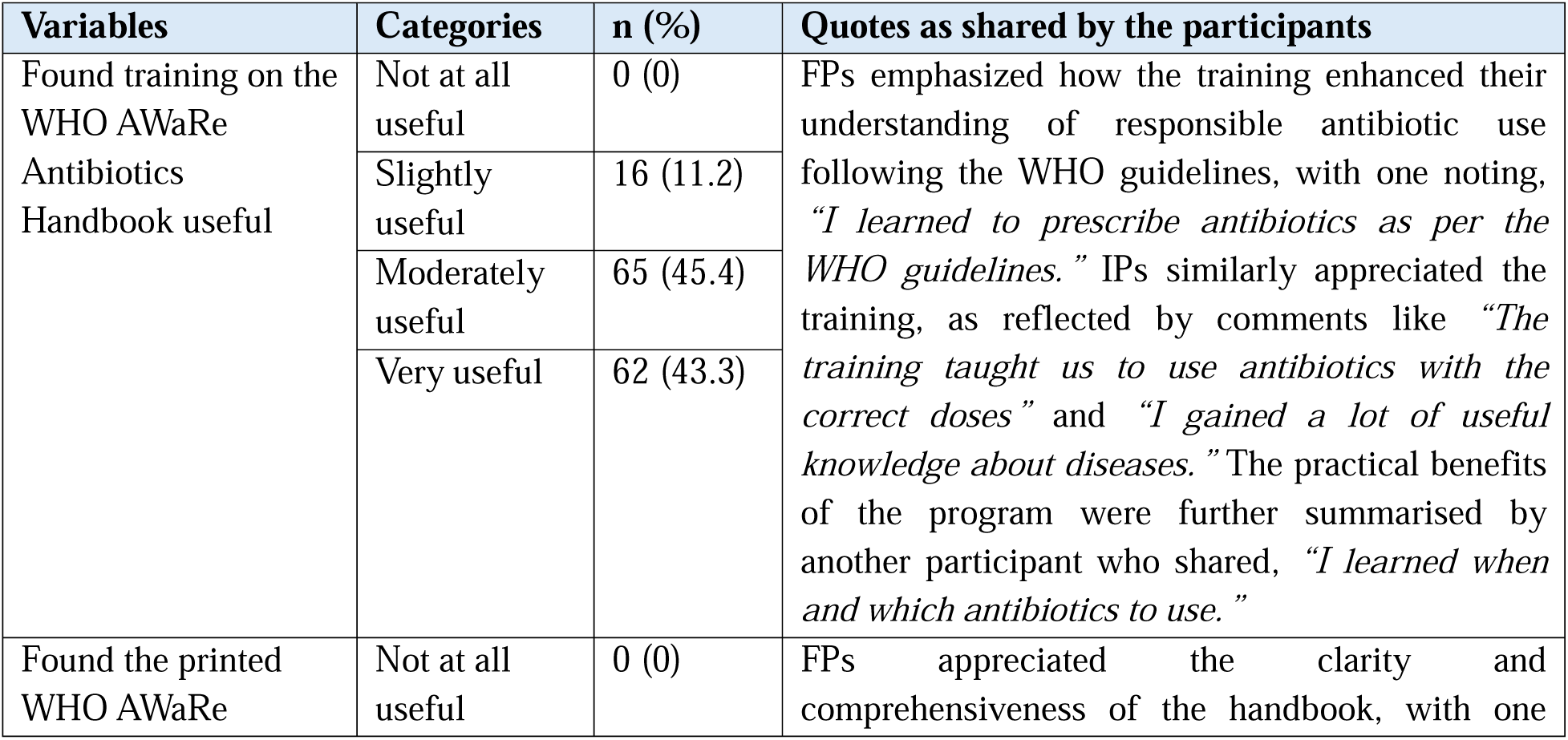
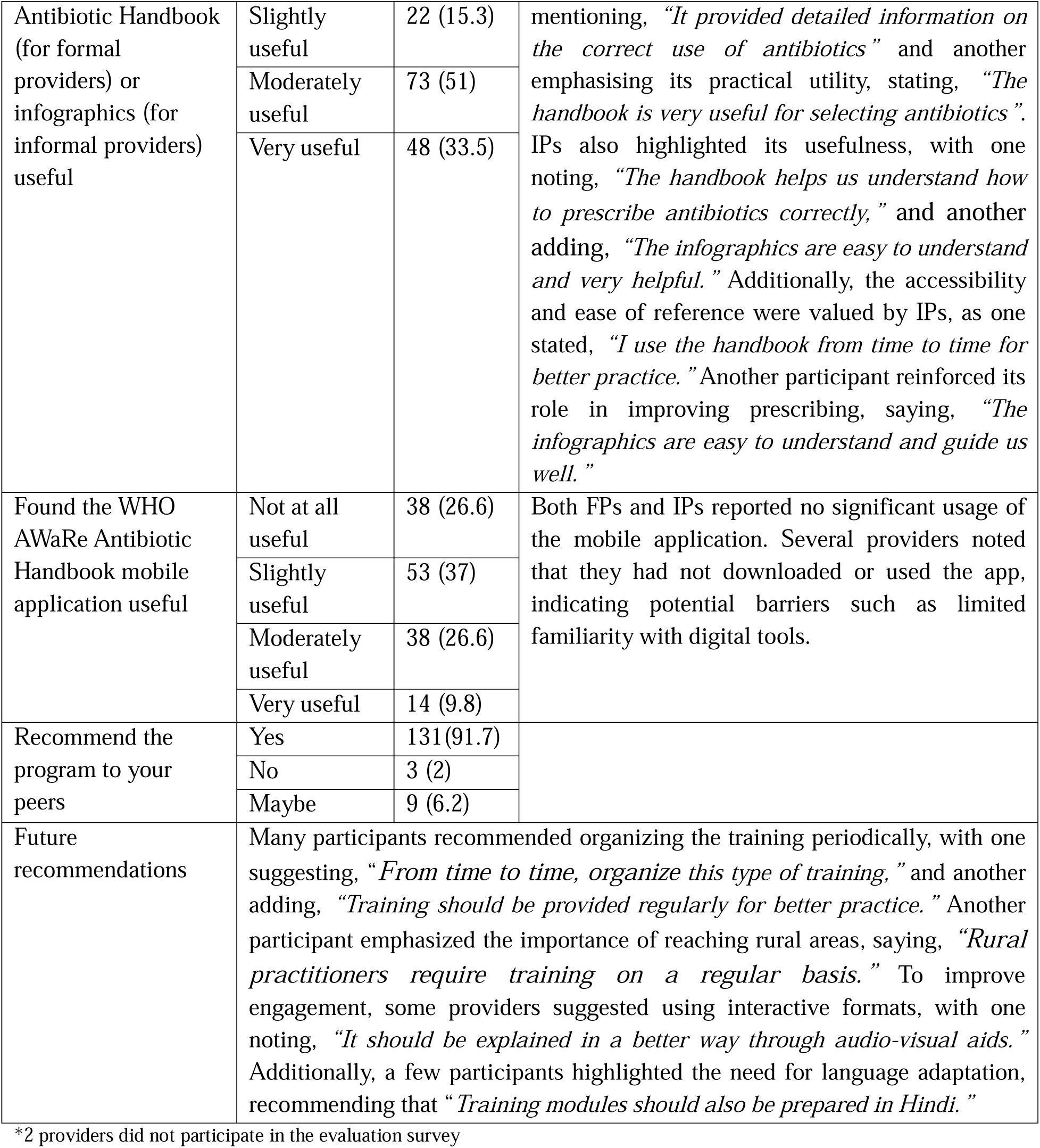
Summary of providers’ feedback on the overall intervention (n=143) *.

In the Watch-category antibiotic prescribing, a significant reduction (p<0.0001) was noted among acute diarrhea cases, with 37 providers stopping and eight starting Watch antibiotic use. No significant changes were observed for cellulitis (p=0.1404), CAP (p=0.1086), or UTI (p=0.1048).

### Change in overall antibiotic prescription and antibiotic prescription in concordance with AWaRe Handbook for four infectious clinical vignettes before and after intervention by provider type

Among IPs (n=105), antibiotic prescribing significantly declined post-intervention for acute diarrhea (p=0.0044) and UTI (p=0.0090), with more providers stopping than starting antibiotic use. No significant changes were observed for cellulitis (p=0.4807) or CAP (p=0.4049). Among FPs (n=40), a reduction in antibiotic prescribing for acute diarrhea was noted post-intervention but was not statistically significant (p=0.0703), and prescribing patterns remained stable for cellulitis (p=1.0000), CAP (p=0.4531), and UTI (p=1.0000).

Regarding antibiotic prescribing in concordance with the WHO AWaRe Handbook, IPs showed a significant improvement post-intervention for acute diarrhea cases (p=0.0044) and a significant decline for cellulitis cases (p=0.0125). A trend towards improvement was also noted for CAP cases (p=0.0987), although it was not statistically significant. Among FPs, no statistically significant improvements were observed across any condition. However, a trend towards improvement was noted for acute diarrhea cases (p=0.0703).

### Participants’ feedback on the overall intervention

The majority of respondents found the intervention useful, with 75% (n=107) rating the training on the AWaRe Handbook as moderately or slightly useful, including 43.3% (n=62) who described it as very useful. Similarly, 84.5% (n=121) found the printed Handbook or infographics useful, with 33.5% (n=48) rating them as very useful. In contrast, the mobile application received mixed feedback; 26.6% (n=38) reported that it was not at all useful, 37% (n=53) found it slightly useful, and only 9.8% (n=14) rated it as very useful. Additionally, 91.7% (n=131) of respondents indicated they would recommend the program to their peers.

## Discussion

Our study provides new insights into the impact of the WHO AWaRe Handbook training in a setting with widespread antibiotic use. The intervention significantly reduced antibiotic prescribing for acute diarrhea and UTI, particularly among IPs. A significant reduction in Watch-category antibiotic use for acute diarrhea was also observed. However, no significant changes were found for cellulitis or CAP, highlighting condition-specific variations in prescribing knowledge.

The lack of change in overall antibiotic prescriptions for cellulitis and CAP cases is expected, as antibiotics are generally indicated for these conditions, and their treatment does not rely on laboratory tests, as outlined in the AWaRe Handbook. A significant decrease in antibiotic prescriptions for UTI cases among IPs is particularly interesting. One possible explanation is that the AWaRe Handbook recommends antibiotic treatment only if there is a compatible clinical presentation and a positive test result (e.g., positive urine leucocytes/leucocyte esterase or positive urine culture). Since our clinical vignettes did not provide urine examination or culture results, some providers may not have prescribed antibiotics immediately, contributing to the reduction in antibiotic prescriptions during the post-intervention phase. We did not systematically collect laboratory diagnostic information for the clinical vignettes, so we could not explore whether confirmatory urine tests increased during the post-intervention phase.

The intervention had mixed effects on the prescription of Watch-category antibiotics. A significant reduction was observed for acute diarrhea, but no significant changes were found for cellulitis, CAP, or UTI. The decrease in Watch-category use for acute diarrhea was primarily driven by reduced overall antibiotic prescribing among IPs. A non-significant declining trend in Watch antibiotic use was observed for CAP, primarily among IPs who shifted from Watch to Access-category antibiotics. For UTI, no significant change in Watch-category prescribing was observed.

The lack of a significant decline in Watch-category use for cellulitis, CAP, and UTI cases highlights the complexity of decision-making regarding antibiotic selection. Several factors may have influenced this trend, including limited familiarity with Access-group antibiotics, lack of awareness of generic names due to reliance on branded medications, and the perception that Access antibiotics are less effective. For instance, a recent study examining the antibiotic stocks of 196 IPs in West Bengal, India, found that none of the IPs stocked nitrofurantoin or trimethoprim-sulfamethoxazole, both of which are WHO-recommended first-line treatments for UTI. ^27^ For cellulitis, Watch-category antibiotic use remained high (among all other conditions) and there was a significant decline in concordance with WHO AWaRe Handbook antibiotic prescribing post-intervention among IPs. One possible explanation is that cellulitis was not explicitly covered during training, unlike the other three conditions. Additionally, acute diarrhea, UTI, and CAP are frequently encountered in primary care settings in India ^28^, meaning providers likely had prior experience managing these cases and may have felt more confident adjusting their prescribing behaviour. In contrast, cellulitis presents diagnostic uncertainty and perceived severity, leading providers, particularly IPs, to favour broad-spectrum antibiotics as a precautionary measure, a pattern reported in previous research. ^29^ ^30^

Overall, the observed reductions in antibiotic prescriptions (for some conditions) align with previous research demonstrating the effectiveness of targeted training interventions in improving primary care providers’ knowledge and behaviour of appropriate antibiotic use. ^31^ Similar trends have been reported in studies specifically focused on IPs. ^32^ ^33^ These findings highlight the potential of training programs while demonstrating the need for condition-specific and sustained strategies to modify antibiotic prescribing effectively.

It is important to note that our study used clinical vignettes to assess changes in prescribing knowledge rather than actual prescribing practices. Vignettes are a well-established tool for evaluating provider decision-making in hypothetical scenarios, but do not fully capture real-world prescribing behaviours, which are influenced by factors such as patient demand, provider characteristics, and healthcare system constraints or incentives. ^29^ ^34^ Therefore, our findings should be interpreted as indicative of improvements in knowledge rather than definitive changes in actual clinical practice. Future studies should complement vignette-based assessments with prescription audits, standardized patient methods, or observational data to assess actual prescribing behaviours.

A key observation from this study was the greater impact of training on IPs, who exhibited significant reductions in antibiotic prescribing for some conditions, such as acute diarrhea (16 reduced versus three increased; p=0.0044), and substantial improvements in prescribing concordant with WHO AWaRe Handbook recommendations for acute diarrhea, while FPs showed smaller, non-significant changes. This difference may be explained by lower baseline awareness of antibiotics among IPs, as reported in surveys ^35^ ^36^, making them more receptive to training intervention. For instance, a randomized controlled trial conducted in West Bengal, India, demonstrated that IPs showed greater improvements in quality of care following a structured training programme. ^9^

The findings on antibiotic prescription in concordance with the WHO AWaRe Handbook indicate only partial alignment. While a change in knowledge is essential to influence antibiotic prescribing, the findings also demonstrate that knowledge alone is insufficient to achieve full adherence to the guidelines. Multiple contextual factors shape providers’ prescribing behaviour, including the availability of Access-category antibiotics, access to diagnostics, patient demand for antibiotics, economic incentives, provider characteristics, pharmaceutical industry influence, and healthcare system constraints. ^30^ ^34^ ^37^ Training combined with audit-feedback mechanisms, ongoing education, peer engagement, regulatory support and enforcement, and patient or community education campaigns has proven effective among PCPs in various settings. ^31^ ^33^ Furthermore, as our intervention was a one-time training delivered to providers, sustained interventions with more extended follow-up periods, refresher training and reinforcement sessions are needed for long-term impact ^31^ ^33^, which was also one of the key recommendations made by the PCPs who participated in our research.

Our results indicate that all IPs (100%) found the training useful to some extent, ranging from slightly to very useful. This finding is particularly encouraging, as a study from India involving multiple key stakeholders has similarly recommended interventions for IPs that promote optimal guidelines for a limited range of antibiotics and healthcare conditions. ^27^ Our study also highlights the importance of adapting the AWaRe Handbook to the local context. Specifically, translating infographics into local languages enhanced accessibility for IPs who are less comfortable with English. While printed materials were well-received, the digital application was less utilized, possibly due to low digital literacy and apprehension among providers toward using a medical application, as reported in other studies. ^38^ Although a digital application was included in our study, the primary emphasis remained on the printed AWaRe Handbook and infographics. Future research should assess the usability of digital applications, explore alternative formats to enhance accessibility, and incorporate strategies to build digital competencies among providers.

## Strength and limitations

This study evaluates the impact of training PCPs (from both formal and informal sectors) on the AWaRe Handbook in India. The use of clinical vignettes allowed for a standardized assessment of prescribing knowledge, and the inclusion of both IPs and FPs provides preliminary, valuable insights into differential training impacts. Additionally, the study was implemented in collaboration with a local NGO, enhancing its real-world applicability and feasibility for scaling similar interventions. However, the study also has several limitations. The use of clinical vignettes enables effective measurement of knowledge, but it does not directly reflect the providers’ real-world prescribing practices. The sample size was relatively small (especially for FPs), which may have reduced statistical power for detecting moderate effects. Furthermore, the 2–3 months follow-up may not have been sufficient to capture long-term changes in antibiotic prescribing.

## Conclusion

This study provides preliminary evidence on the effectiveness of training PCPs using the WHO AWaRe Antibiotic Handbook to improve antibiotic prescribing knowledge. The intervention significantly improved prescribing knowledge for acute diarrhea, with a greater impact among IPs, highlighting their responsiveness to structured training. The majority of providers rated the training as moderately or very useful. While these findings support the potential of the WHO AWaRe Antibiotic Handbook as an intervention, they also highlight the need for tailored approaches and sustained engagement to achieve long-term improvements in antibiotic prescribing. Future research should focus on longer follow-up periods, larger randomized controlled trials, and real-world prescribing data to evaluate the long-term impact and scalability of such interventions in LMICs’ primary care settings.

## Contributors

MP and SAR led the funding acquisition. MP, SAR, SG, PT, PS, CJ, and SS contributed to the conceptualization and methodology. Field implementation was carried out by PS, CJ, PT, and SG. Data collection and supervision were undertaken by PKS, PS, CJ, PT, and SG. PT, SG, CJ, SS, DT, and MD were responsible for data management and analysis. PT and SG wrote the original draft. PT, PS, CJ, SS, PKS, DT, MD, MP, SAR, and SG contributed to manuscript review and approval. MP and SAR are the guarantors of the study.

## Data availability statement

The data underlying the study’s results are available in an anonymized format upon reasonable request by emailing the corresponding author at.

## Competing interests

The authors have declared that no competing interests exist.

## Ethics approval and consent to participate

Prior to the commencement of training and data collection activities, written informed consent was obtained from all enrolled providers by trained field officers. The consent process involved providing participants with detailed information about the study’s objectives, procedures, potential risks and benefits, and their rights as research participants, including the right to withdraw at any time without any consequences. The information was conveyed in a language preferred by the participants, and consent was documented through participant signatures.

This study received ethical approval from the Research Ethics Board of the Faculty of Medicine and Health Sciences at McGill University (Protocol Number: A08-E32-23B) and the McGill University Health Centre (MUHC) Ethics Board (Protocol Number: MI4 SP AMR/2024-10007). All research activities were conducted in accordance with the ethical principles outlined in the Declaration of Helsinki and relevant national guidelines on research involving human participants.

## Funding

This study was supported by grant #SFG5-01, awarded by the McGill Interdisciplinary Initiative in Infection and Immunity (MI4) on May 1st, 2023. The study’s funder had no role in study design, data collection, analysis, interpretation, or manuscript writing.

## Acknowledgements

We wish to thank all the field officers from World Health Partners who were involved in data collection. Our special thanks to all the primary care providers who participated in this research.

## Notes

### Competing Interest Statement

The authors have declared no competing interest.

### Clinical Trial

Not registered. It is an interventional research but carried out as a pilot pre-post study

### Author Declarations

This study received ethical approval from the Research Ethics Board of the Faculty of Medicine and Health Sciences at McGill University (Protocol Number: A08-E32-23B) and the McGill University Health Centre (MUHC) Ethics Board (Protocol Number: MI4 SP AMR/2024-10007).

